# *LMNA* R644C associates with hepatic steatosis in a large cohort and increases cellular lipid droplet accumulation *in vitro*

**DOI:** 10.1101/2023.12.20.23300290

**Authors:** Kapil K. Upadhyay, Xiaomeng Du, Yanhua Chen, Elizabeth K. Speliotes, Graham F. Brady

## Abstract

The R644C variant of lamin A is controversial, as it has been linked to multiple phenotypes in familial studies, but has also been identified in apparently healthy volunteers. Here we present data from a large midwestern US cohort showing that this variant associates genetically with hepatic steatosis, and with related traits in additional publicly available datasets, while *in vitro* testing demonstrated that this variant increased cellular lipid droplet accumulation. Taken together, these data support this *LMNA* variant’s potential pathogenicity in lipodystrophy and metabolic liver disease.

---

Metabolic dysfunction-associated steatotic liver disease (MASLD) and, when associated with lipotoxicity and inflammation, metabolic dysfunction-associated steatohepatitis (MASH) together represent a genetically and phenotypically diverse entity that is now the most common liver disease in the United States and for which there is no approved medical therapy^1,2^. Therefore, it is imperative to define the full spectrum of its pathogenesis to facilitate both the development of novel therapies and their delivery to those who are most likely to benefit. A novel and relatively under-explored area in MASLD/MASH is the role of the nuclear envelope and lamina; variants in *LMNA*, encoding A-type nuclear lamins, are implicated in diverse diseases including progeria, muscular dystrophy, and lipodystrophy syndromes that include insulin resistance and early-onset MASH with progression to cirrhosis^3,4^. Therefore, a fuller understanding of nuclear lamina-related liver disease may provide valuable insights into MASH generally; however, the mechanisms of *LMNA*-related liver disease are largely obscure, and some *LMNA* variants exhibit significant variability in phenotype and penetrance within and between families. The *LMNA* R644C variant, which alters proteolytic processing of lamin A^5^, has been controversial and appears to be particularly variable, with reported linkages to several distinct phenotypes but also identification in healthy volunteers^6,7^. Therefore, we sought to clarify the potential pathogenicity of this variant in MASLD by determining its genetic association with hepatic steatosis in a large single-center US cohort and its functional effects on lipid droplet accumulation *in vitro*.

We tested rs142000963 (g.156138719 C>T; *LMNA* p.R644C) for its effect on hepatic steatosis in the Michigan Genomics Initiative (MGI) cohort (>57,000 individuals)^8,9^. Natural language processing of pathology and radiology reports was used to identify cases with hepatic steatosis (n=5,856) on liver biopsy and/or imaging; participants not classified as cases were considered controls (n=51,166). All MGI participants had been genotyped via the Illumina HumanCoreExome array; overall rs142000963 minor allele frequency (rs142000963-T) was 0.002. Association analysis was performed in SAIGE v0.29 with steatosis as the outcome, controlling for age, age^2^, sex, and the first 10 principal components in an additive genetic model. We found that *LMNA* R644C positively associated with hepatic steatosis, with an odds ratio of 1.7 (*P*=0.02; **Table 1**). The strength and significance of the association did not vary between the all-ancestry MGI cohort (N=57,022) and the European ancestry-only cohort (n=51,550). To determine whether rs142000963-T might predispose to more advanced liver disease in addition to hepatic steatosis, a phenome-wide association study (PheWAS) was performed in the MGI dataset using rs142000963-T as the variant of interest. We found that rs142000963-T significantly associated with hepatic decompensation – development of ascites (*P*=0.002, odds ratio = 5.0; **Supplementary Table 1**) – which remained significant after Benjamini-Hochberg (with false-discovery rate of 0.05) or Bonferroni correction for simultaneous testing (all liver-related phenotypes listed in **Supplementary Table 2)**. Weaker associations, which were not significant after Benjamini-Hochberg correction, were seen with undergoing liver transplant (*P*=0.03, odds ratio = 10.0), and acute or subacute hepatic necrosis (*P*<0.05, odds ratio = 16.4). Consistent with its proposed role as an atypical or incompletely penetrant lipodystrophy allele^7^, PheWAS of publicly available data from larger datasets via the Type 2 Diabetes Knowledge Portal^10^ (T2DKP, https://t2d.hugeamp.org/) revealed strong associations between rs142000963-T and extrahepatic MASLD/MASH-related anthropometric, glycemic, and lipid-related traits including waist-to-hip ratio (*P*=0.001), type 2 diabetes (*P*=0.004), higher hemoglobin A1c (*P*=0.001), and decreased HDL (*P*=0.004); **Supplementary Table 3**. These associations remained significant after Benjamini-Hochberg correction for 45 such phenotypes with false-discovery rate of 0.05; tested phenotypes are listed in **Supplementary Table 4**.

**Table 1.**
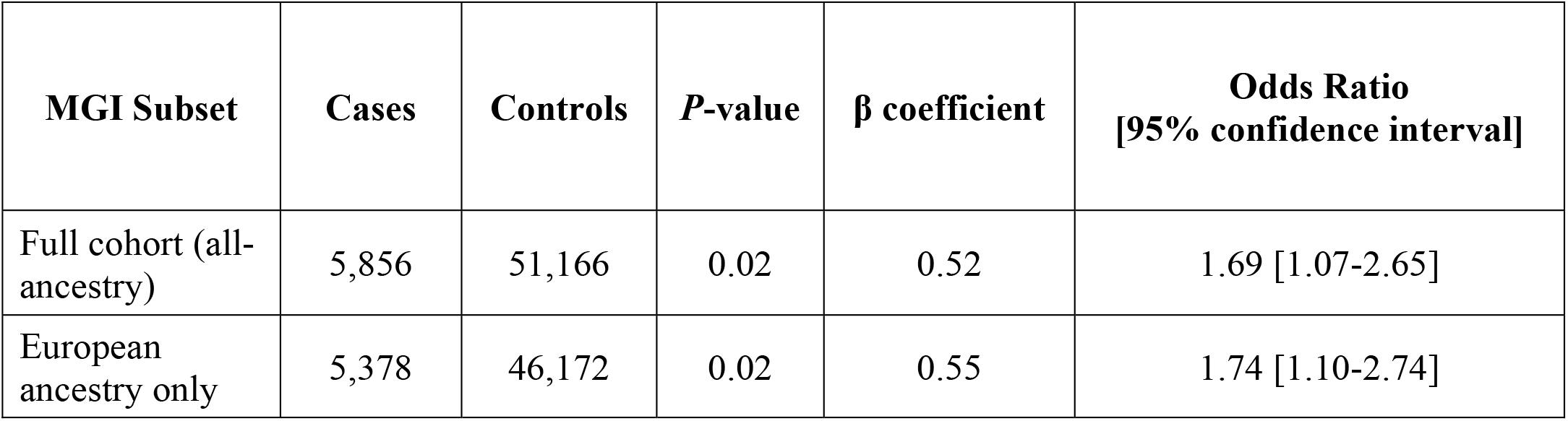
Association of rs142000963-T (*LMNA* R644C) with hepatic steatosis in MGI. Positive association of rs142000963-T with hepatic steatosis was equally strong within the all-ancestry analysis (top row) and the self-reported European ancestry subset of MGI (bottom row).

The R644C variant of lamin A has been shown *in vitro* to alter its proteolytic processing by the zinc-dependent protease ZMPSTE24, but the functional impact of this altered processing has been unclear, as this variant has been linked to disparate laminopathy phenotypes^5-7^. Given the results of our GWAS and PheWAS analyses, we sought to address whether the steatosis-promoting effects of rs142000963-T could be hepatocyte-autonomous. To address this, mCherry-tagged wild-type (WT) or R644C lamin A was expressed in Huh7 human hepatoma cells, and lipid accumulation was determined by fluorescence microscopy with a lipid-binding fluorophore. Relative to WT lamin A, cells expressing lamin A R644C demonstrated significantly increased lipid droplet accumulation, without (**Figure 1A**) or with (**Figure 1B**) oleic acid supplementation (quantitation shown in **Figure 1C**). These functional data corroborate our genetic data and support the pathogenicity of rs142000963-T in *LMNA*-related lipodystrophy and MASLD/MASH; in addition, they suggest the possibility of hepatocyte-autonomous lipid accumulation *in vivo*.

**Figure 1.**
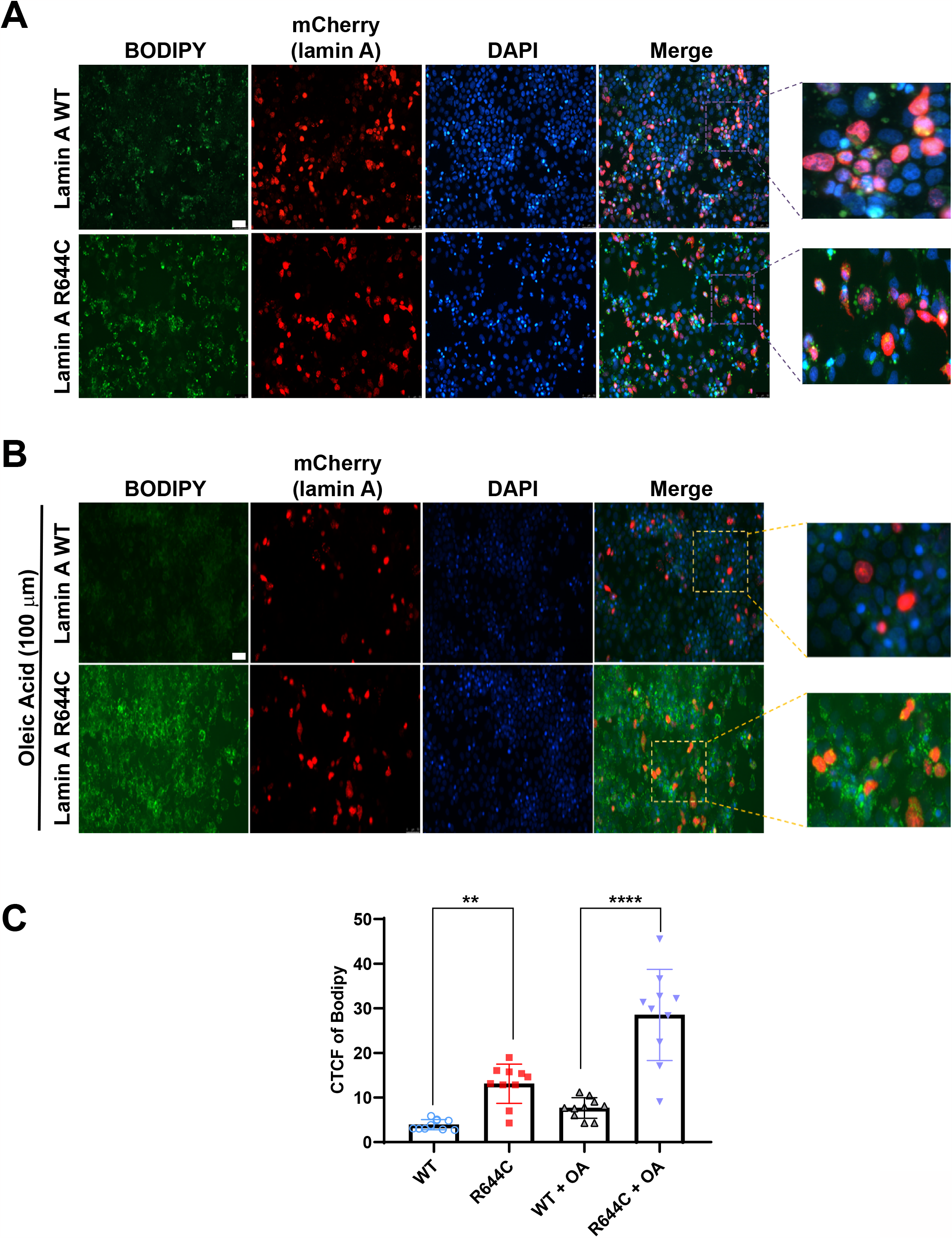
Lamin A R644C increased lipid accumulation in transfected human hepatoma cells. ***A***. Huh 7 cells were transfected with mCherry-tagged WT or R644C lamin A, then fixed and stained for lipid droplet accumulation with BODIPY 493/593 after 24 h; results shown are representative of n=3 experiments. ***B***. Huh 7 cells were transfected with mCherry-tagged WT or R644C lamin A for 24h, followed by overnight treatment with oleic acid (OA; 100 μM) in serum-free medium and post-fixation lipid staining with BODIPY 493/593; results shown are representative of n=3 experiments. In both ***A*** and ***B***, WT and R644C lamin A-transfected cells were imaged at the same time with the same exposure settings; however, images shown in ***A*** (no lipid treatment) and ***B*** (oleic acid treatment) were obtained separately and thus cannot be directly compared. Scale bars = 50 μm. ***C***. Lipid staining data shown in ***A*** and ***B*** were quantitated with ImageJ, and results are expressed as mean ± S.D. \*\**P*<0.01; \*\*\*\**P*<0.0001.

In summary, rs142000963-T (*LMNA* R644C) significantly associated with hepatic steatosis, and to a lesser extent with liver-related events in a large midwestern US cohort, as well as with MASLD-related metabolic traits in large publicly available datasets via T2DKP; moreover, it increased lipid droplet accumulation in Huh7 cells. These data provide genetic support, and direct functional evidence, for the pathogenicity of rs142000963-T / *LMNA* R644C in metabolic laminopathies and suggest that its incomplete penetrance may be due, at least in part, to genetic modifiers that have not yet been defined.

## Data Availability

All relevant data are included within the manuscript and its accompanying supplementary material or are publicly available.

## Notes

**Grant Support:** This work was supported by K08DK120948 (G.F.B.) and the University of Michigan Department of Internal Medicine. X.D., Y.C., and E.K.S. are supported in part by R01DK106621 (E.K.S.), R01DK107904 (E.K.S.), R01DK128871 (E.K.S.), R01DK131787 (E.K.S.), the University of Michigan Department of Internal Medicine, and a University of Michigan MBioFAR award.

### Competing Interest Statement

The authors have declared no competing interest.

### Funding Statement

This work was supported by K08DK120948 (G.F.B.) and the University of Michigan Department of Internal Medicine. X.D., Y.C., and E.K.S. are supported in part by R01DK106621 (E.K.S.), R01DK107904 (E.K.S.), R01DK128871 (E.K.S.), R01DK131787 (E.K.S.), the University of Michigan Department of Internal Medicine, and a University of Michigan MBioFAR award.

### Author Declarations

IRB of the University of Michigan gave ethical approval for this work.

