## Supplementary Material for "*LMNA* R644C associates with hepatic steatosis in a large cohort and increases cellular lipid droplet accumulation *in vitro*"

**Table S1**. Liver-related phenotypes associated with rs142000963-T in MGI.

| Phenotype | Direction | *P*-value | Odds ratio [95% confidence interval] |
|---|---|---|---|
| Non-malignant ascites | Positive | 0.002 | 5.0 [1.9-13.2] |
| Liver replaced by transplant | Positive | 0.033 | 10.0 [1.2-86.1] |
| Acute and subacute necrosis of liver | Positive | 0.046 | 16.4 [1.1-255.7] |

**Table S3**. Association of rs142000963-T with extra-hepatic metabolic phenotypes (via the Type 2 Diabetes Knowledge Portal).

| Phenotype | Direction | *P*-value |
|---|---|---|
| Waist to hip ratio | Positive | 0.001 |
| Type 2 diabetes | Positive | 0.004 |
| Hemoglobin A1c | Positive | 0.001 |
| HDL | Negative | 0.004 |

### Supplementary Methods

***Cohort and Ethics Statement.*** Michigan Genomics Initiative (MGI) participants provided written informed consent approved by the University of Michigan IRB, which approved all human subjects research in this study.

***Association analysis.*** We tested rs142000963 (GRCh38 g.156138719C>T; *LMNA* p.R644C) for effects on hepatic steatosis in the MGI cohort (freeze 3; N=57,022 individuals). Natural language processing of pathology and radiology reports was used to identify cases (n=5,856) with hepatic steatosis on liver biopsy or imaging; participants not classified as cases were considered controls (n=51,166). All included MGI participants had been genotyped via the Illumina HumanCoreExome array, which includes rs142000963; overall minor allele frequency was 0.002. Association analysis was performed in SAIGE v0.29 with steatosis as the outcome, controlling for age, $age^2$, sex, and the first 10 principal components in an additive genetic model.

***Phenome-wide association study (PheWAS).*** PheWAS was performed for *LMNA* R644C (rs142000963-T) in MGI freeze 3 using a web-based browser (http://pheweb.org; last accessed 12/4/2023), with a focus on liver-related phenotypes. Liver-related phenotypes that significantly associated with rs142000963 ($P<0.05$) were included Supplementary Table 1. PheWAS of extra-hepatic metabolic traits was performed with publicly available data via the Type 2 Diabetes Knowledge Portal (T2DKP, https://t2d.hugeamp.org/ or type2diabetesgenetics.org, RRID SCR_003743; last accessed 12/11/2023)[1].

***Cell culture and treatment conditions.*** Huh7 cells were maintained in DMEM containing glucose and lacking L-glutamine, supplemented with 10% heat-inactivated fetal bovine serum (Thermo Fisher A3840202, USA) at 37°C, 5% $CO_2$. For transfection, cells were plated at ~60-70% confluency one day prior to transfection. For lipid staining, cells were plated in 4-well chamber slides in DMEM with 10% FBS, then transfected with mCherry-lamin A the next day. For lipid supplementation, culture medium was changed 24 hours post-transfection to DMEM containing 1% fatty acid–free bovine serum albumin (Sigma-Aldrich) and oleic acid (100 μM; Sigma-Aldrich), and cells were incubated for an additional 24 hours. For staining, cells were fixed in 4% paraformaldehyde (PFA) in PBS for 15 mins at 22°C, then washed in PBS; neutral lipids were stained with 10 μM boron dipyrromethene 493/503 (BODIPY; Thermo Fisher). Quantification of green BODIPY fluorescence was performed using ImageJ version 1.53t for at least three 20× fields per condition per experiment and expressed as total BODIPY fluorescence per field (CTCF) which was calculated as CTCF = Integrated Density – (Area of selected cells × Mean fluorescence of background readings).

***Plasmids and transfection.*** Amino-terminally tagged human *LMNA* was described previously[2]. The *LMNA* open reading frame was cloned into mCherry-Parkin, Addgene plasmid #23956 (replacing the Parkin open reading frame), via the BspEI and BamHI sites. mCherry-Parkin was a gift from Richard Youle (Addgene plasmid #23956; http://n2t.net/addgene:23956; RRID:Addgene_23956)[3]. The R644C variant of *LMNA* was generated using the QuikChange II site-directed mutagenesis kit (Agilent Technologies, Santa Clara, CA). Lipofectamine-2000 (Life Technologies, Carlsbad, CA) was used for transfection of Huh7 cells according to the manufacturer's instructions.

***Statistical analysis.*** For PheWAS testing, the Benjamini-Hochberg method was used to correct for simultaneous testing of multiple phenotypes, with false-discovery rate set to 0.05. Statistical analysis of *in vitro* data was performed in GraphPad Prism v.8. To compare >2 groups (Figure 1C), 1-way ANOVA was performed in Graphpad followed by Tukey's multiple comparison test.
